# The NeuroBioBank Whole-Genome Catalog: Sequencing from human brain donors with central nervous system disorders

**DOI:** 10.1101/2024.08.29.24312734

**Authors:** Daniel Hupalo, Jacob L. McCauley, Lissette Gomez, Anthony J. Griswold, Gabriela Hoher, Ioanna Konidari, Jose Lorenzo, Griffin S. Parker, Julianna Pascual, Amanda R. Sandford, Patrice L. Whitehead, David A. Davis, Susanna Garamszegi, S. Humayun Gultekin, Xiaoyan Sun, Regina T. Vontell, Michael Chatigny, Darren Chernicky, Myrtha M. Constant, Isabelle G. Darling, David J. Ennulat, John M. Esposito, Kiely Morris, Elisabeth S. Lawton, Neda R. Morakabati, Phyllis Oduor, Allison P. Rodgers, Lorelle A. Sang, Kathleen M. Sullivan, Catalina J. Tabit, Tori Turpin, Aya Zeabi, Tina Zheng, Sabina Berretta, Torsten Klengel, Derek H. Oakley, W. Brad Ruzicka, Thomas Blanchard, Eric Ho, Robert Johnson, Alexandra LeFevre, Maxwell Bustamante, Vahram Haroutunian, Pavel Katsel, Christine Marino, Stephen Panopoulos, Dushyant P. Purohit, Michael Wysocki, Jill R. Glausier, David .A. Lewis, Rashed M. Nagra, Camille Alba, Julianna Martin, Elizabeth Rice, John Rosenberger, Grace Smith, Gauthaman Sukumar, Miranda Tompkins, Matthew Wilkerson, Clifton L. Dalgard, William K. Scott

**Affiliations:** The American Genome Center, Center for Military Precision Health, and Department of Anatomy, Physiology and Genetics, Uniformed Services University, Bethesda, MD; John P. Hussman Institute for Human Genomics, Miller School of Medicine, University of Miami, Miami, FL; Brain Endowment Bank, Department of Neurology, Miller School of Medicine, University of Miami, Miami, FL; Henry M. Jackson Foundation for the Advancement of Military Medicine, Rockville, MD; Department of Psychiatry, Icahn School of Medicine at Mount Sinai, New York, NY; Department of Pathology, Icahn School of Medicine at Mount Sinai, New York, NY; Friedman Brain Institute, Icahn School of Medicine at Mount Sinai, New York, NY; Department of Neuroscience, Icahn School of Medicine at Mount Sinai, New York, NY; Mental Illness Research Education and Clinical Center (VISN 2 South), James J. Peters VA Medical Center, Bronx, NY; McLean Hospital, Belmont, MA; Harvard Medical School, Boston, MA; Broad Institute of MIT and Harvard, Cambridge, MA; University of Maryland Brain and Tissue Bank, University of Maryland Medical School, Baltimore, MD; Department of Psychiatry, University of Pittsburgh School of Medicine, Pittsburgh, PA; Human Brain and Spinal Fluid Resource Center, Brentwood Biomedical Research Institute, Los Angeles, CA

**Author notes:** Co-first author (Daniel Hupalo, Jacob L. McCauley).

## Abstract

Central nervous system diseases are a prevailing cause of morbidity and mortality worldwide, and are influenced by environmental and biological factors including genetic risk. Here we generated genome-wide genetic data on a large cohort of brain tissue donors with in-depth clinical and neuropathological phenotyping, allowing for broad investigations into the risk and mechanisms of these neurological, neurodevelopmental, and psychiatric conditions. This resource consists of 9,663 donors with array-based genotyping and 9,543 donors with whole-genome sequencing completed. The clinical diagnoses of these donors include 148 central nervous system diseases clustered in 15 broad categories by ICD-10 coding. These donors were collected by six repositories comprising the NIH NeuroBioBank, with an average participant age of 60 years. While primarily older individuals of European descent, the cohort also contains younger donors and individuals from non-European backgrounds. Variants detected by Whole-Genome Sequencing (WGS) were called genome-wide and annotated to describe their functional impact, resulting in 171,121,209 unique mutations and 1,078,774 non-silent mutations. This whole-genome resource has been made available in the NIMH Data Archive (nda.nih.gov) and accompanying deep demographic and phenotypic descriptions are available at the NeuroBioBank Portal (neurobiobank.nih.gov). To illustrate an application of this resource, we replicated the strong association observed in previous studies between pathogenic CAG repeat expansions in the *HTT* gene with the clinical diagnosis of Huntington’s disease.

## INTRODUCTION

Biorepositories are a vital resource for cross-sectional and longitudinal studies of diseases (1). The National Institutes of Health NeuroBioBank (NIH NBB) is a broad effort to collect central nervous system tissue from a diverse spectrum of neurological and psychiatric diseases, to carefully process, characterize and store these biospecimens, and to distribute samples for use in biomedical research studies across the world (2). The NBB operates a federated model involving six biobank locations to maximize geographic coverage and sample quantity while preserving sample quality through common protocols and procedures (3). Together, these locations collect tissue samples and clinical data that contribute to a central collection for use in scientific research.

Central nervous system illnesses affect a large number of individuals worldwide and are intrinsically linked to many environmental and biological factors (4, 5). Association studies utilizing genotype, exome- and whole-genome sequencing datasets have explored the genetics of specific diseases and identified many common and few rare genetic variants that contribute to their development (6). Advancing our collective knowledge of neurological and psychiatric diseases will require large sample sizes, diverse populations of individuals, and deep phenotyping of disease among patients to identify variation which may contribute to their development (7-9). By embarking on a deep cataloging of rare variation using whole-genome sequence paired with bio-banked tissue, we can extend the characterization of each of these central nervous system diseases (10)

Cohorts with large sample sizes power hypothesis-driven investigations, and it is the mission of tissue repositories to curate these large collections of post-mortem central nervous system tissue (11). As tissue requests are increasing to conduct large-scale -omic analyses of brain tissue, it has become desirable to pro-actively annotate these brain samples with genome-wide genotyping or sequencing data as a resource for the scientific community (12). Initiatives such as this study provide an efficient means for providing germline genomic variation to a wide variety of researchers, as the genomic variation needs only to be assessed one time and then be made available with the tissue specimen. Here, we conduct genome-wide genotyping and sequencing of samples within the NBB cohort, collected prior to the end of 2021, to add genetic annotation to the extensive neuropathologic and clinical annotation available on these brain tissue donors, thus increasing the utility of this resource to the scientific community. This manuscript describes the selection of samples, generation of called genotypes from whole-genome sequencing, integration of genomic and phenotypic annotation, and illustrates utility through analysis of an exemplar monogenic disease with a known genetic basis, Huntington’s disease.

## METHODS

### Subjects, phenotypes, brain tissue sampling and DNA extraction

The six NIH NeuroBioBank sites include the University of Miami Brain Endowment Bank, the Harvard Brain Tissue Resource Center, The Human Brain and Spinal Fluid Resource Center (Sepulveda), the Mount Sinai NeuroBioBank, the University of Maryland Brain and Tissue Bank, and the Brain Tissue Donation Program at the University of Pittsburgh. These Brain Tissue Repositories (BTRs) reviewed existing collections and identified 10,270 frozen tissue samples with sufficient quantity and neuropathologic/clinical annotation for genotyping and whole-genome sequencing. All samples were obtained from individuals with appropriate consent from the local institutional review boards. While each NeuroBioBank BTR follows local protocols for clinical and neuropathological assessment ante- and post-mortem, in general, phenotyping of both kinds was conducted retrospectively through semi-structured interviews of knowledgeable informants, review of available medical records, and neuropathological assessment of brain tissue.

Within the publicly accessible NIH NBB phenotype database, diagnoses are organized into four different categories: Clinical Brain, Neuropathology, Genetic and Non-Brain. The data field of “Clinical brain diagnosis” reports consensus psychiatric and neurologic diagnoses made based on review of clinical data, including medical records, interview or questionnaires administered to knowledgeable informants, and/or self-reported diagnoses at registration. The data field of “Neuropathology diagnosis” reports diagnoses made based on the results of neuropathological examination by a qualified neuropathologist. The data field of “Genetic diagnosis” reports diagnoses made based on genetic testing. The data field of “Non-brain diagnosis” reports clinical diagnoses that do not primarily affect brain structure or function, but could have some effects on the brain. Because BTRs follow local protocols for clinical brain diagnostic procedures, the level of evidence used to determine a clinical brain diagnosis is captured with the data field “Basis of clinical brain diagnosis.” A basis of “Confirmed” reflects that sufficient evidence existed to 1) make a diagnosis based on medical history review, 2) make a diagnosis by the consensus of expert clinicians, and/or 3) confirm the absence of a clinical diagnosis. A basis of “Investigator impressions” reflects that the diagnosis is based on the clinical impressions of the clinician(s) reviewing the available information, but that information was insufficient to refer a Confirmed diagnosis. The data field “Insufficient data” reflects that the available information was insufficient to form a reasonable impression of, or to exclude, any clinical brain-related diagnoses. Subjects may have multiple diagnoses of unrelated diseases within and across diagnostic categories.

A diagnosis may meet criteria for multiple categorizations. Individuals that had no clinical brain diagnosis are indicated with a custom NBB000 “no clinical brain diagnosis found” code, and subjects that had no neuropathological diagnosis are indicated with a custom NBB222 “diagnostic pathology not present” code. When diagnostic information was not captured effectively by ICD-10 coding, the NBB generated additional custom labels. Other custom codes included in the phenotype database describe categories of missing or pending information. Finally, due to the small numbers of individuals of advanced age, all participants with an age >89 were further de-identified by setting the field of age to 89+.

At each BTR, flash frozen tissue aliquots were dissected and maintained at -80°C until DNA extraction. For all BTRs, samples of frozen brain tissue were primarily taken from the occipital pole, Brodmann area 17 (BA17), but other areas such as frontal pole (BA10) or cerebellum were used when BA17 was not available. High molecular weight DNA was extracted from approximately 60-80 mg of tissue using the QIASymphony DSP DNA Midi Kit on the QiaSymphony SP platform following manufacturer’s standard protocols. DNA concentrations were determined using spectrophotometry and extraction quality was measured using agarose gel analysis, using 0.8% agarose gels and ∼0.5ug of DNA, to score Extraction Quality (EQ) on a scale of 0 to 5. A total of 245 samples were removed from the cohort prior to DNA extraction. DNA was successfully obtained from 10,025 brain tissue samples, with each passing sample having an EQ score of 3 or better and DNA quantity >= 2 ug.

### Genotype analysis

DNA from samples meeting quality and quantity metrics underwent genotyping at the HIHG Center for Genome Technology. Samples were organized on 96-well plates and processed in minimum batches of 96 samples. 200ng of DNA from each sample was normalized to 50ng/ul and used for genome-wide genotyping using Illumina’s Infinium Global Screening Array-24 v3.0 (GSAv3) that interrogates approximately 650,000 markers. Each sample preparation followed the Infinium HTS Assay procedures. In brief, 200ng of DNA per sample was amplified, fragmented, and hybridized to a beadchip. The beadchips were then scanned on the Illumina iScan System and processed using the Illumina GenomeStudio v2.0.0 software package. Genotypes were called using the Illumina Genome Studio software and the Illumina iScan System. Samples with call rates below 98% were excluded from analysis and a GenCall cutoff score of 0.15 was used for all Infinium II products. Genotype calls were exported to VCF format from Illumina GenomeStudio for further analysis.

Genotypes were examined for: concordance with biological sex provided by each site, genotype call rate across all markers ≥98%, pairwise genotype matching across samples indicating potential sample duplications and sample swaps. Samples with mismatched reported biological sex (*n*=172), duplication with another genotyped sample (*n*=81), low (<98%) genotype call rate (*n*=68), or other technical issue (*n*=24) were removed from analysis. A total of 9,680 samples were considered of sufficient quality for attempting library preparation for whole-genome sequencing, including 17 related individuals which were later struck from inclusion. Genotyping results were successfully generated for 9,663 samples and were combined into a VCF file which is available to researchers through the NDA portal (nda.nih.gov, collection number 3917, experiment ID 2206).

### Whole-genome sequencing of 9,680 individuals

A total of 9,680 extracted DNA samples contained sufficient quantity and quality of DNA to conduct WGS and samples were shipped at -80 °C to The American Genome Center at Uniformed Services University in Bethesda Maryland on 96-well plates, with one sample per well. Per-plate quality was assessed by checking visual plate integrity for unsealed wells, and DNA quantification was performed and compared to the manifest for agreement. All passing samples were required to have an extracted DNA concentration of at least 30ng/μl. Sequencing libraries were prepared from genomic DNA using 9,680 samples and sequenced using PCR-free tagmentation-based library preparation as previously described (13, 14). Whole-genome sequencing was conducted on the NovaSeq 6000 (Illumina) System, during the data generation period of this study. Sequencing libraries were generated using unique dual indexes for multiplex pooling of 24-32 libraries for sequencing on the NovaSeq 6000 System using the S4 flowcell with 300 cycle SBS kits. All sequencing runs conducted generated 2×150bp reads. A mean coverage threshold of >=30x coverage was used for each unique sample. Sequencing libraries that did not meet this threshold coverage during the first pooled sequencing were re-queued for additional sequencing and data was merged until 30x genome coverage was reached, or excluded from the study if the threshold was not met.

### Summary statistics and variant calling for 9,543 passing genomes

A total of 9,543 unique samples completed sequencing and met all coverage and quality metrics. Reads from individual flowcells were demultiplexed using bcl2FASTQ 2.20 and converted to ORA compressed files using DRAGEN Server 4.0. The resulting sequencing reads were processed using Illumina DRAGEN (Dynamic Read Analysis for Genomics) 4.0 software, a specialized pipeline tool designed for rapid and accurate processing of genomic data and identification of single nucleotide variants (SNVs), small insertions/deletions (INDELs), and structural variants from Illumina sequencers (15-17). DRAGEN alignments and variant calling outputs are cross-compatible with the Genome Analysis Tool Kit (GATK). Default DRAGEN parameters, pre-tuned to human genome alignment and variant calling, were used for mapping, base quality score recalibration, and variant calling.

Each sample was individually aligned using Illumina’s DRAGEN pipeline version 4.0 to produce a CRAM alignment file, and alignment quality statistics. The subsequent CRAM was used for variant calls (SNVs, INDELs, CNVs, and SVs), using the DRAGEN server pipeline as described above. This pipeline produced 16 files for each sample, including the alignment CRAM file, genome VCF, VCF, SV VCF, and their associated metrics, indexes and checksums. Alignment was performed using the GRCh38 genome reference including decoys and alternative haplotypes (ALT contigs) optimized for DRAGEN v4.0, with ALT-aware read mapping. Individual variant call files were post-processed using Illumina’s default Germline Variant Small Hard Filtering parameters.

Using variants from the genotyping analysis described above, 163,249 variants (25% of the GSA variants) were randomly selected from the set of 9,663 genotyped individuals. A subset of 34,328 independent variants (R^2^ < 0.5), from the set of 163,249 variants, on autosomal chromosomes, with genotyping call rate > 90%, and minor allele frequency > 0.2 was then used to calculate concordance between genotype and WGS data for the overlapping 9,520 samples between the genotyping and WGS.

### NeuroBioBank Cohort VCF creation

Whole-genome sequenced samples were evaluated for quality using several metrics. A coverage threshold of >=30x mean genome coverage per sample was used, as calculated by the statistic “Average alignment coverage over genome” as listed in the DRAGEN 4.0 statistics. Contamination per sample was evaluated using VerifyBamID (18). Duplicate or related samples were identified by calculating genomic similarity using a binary distance matrix between variants detected within samples. Biological sex was inferred by coverage of the X and Y chromosomes and matched the self-reported assigned sex at birth in all samples.

Genome-wide short nucleotide variants, including single-nucleotide variants and short insertions and deletions, were combined using the DRAGEN implementation of Illumina’s gvcf-genotyper software, packaged within the DRAGEN 4.0 software suite, which merged individual genome VCF files into a combined ‘cohort VCF’. Loci with multiple different alleles, Multi Nucleotide Polymorphisms (MNPs), were split into Single Nucleotide Polymorphisms (SNPs) in every case, with one line per allele to disambiguate subsequent filtering and annotation steps. Individual gVCFs were joined into gene-aware chunks of up to 10Mb in size, spanning all autosomes, X and Y chromosomes, and the mitochondrial genome (M). Splits between chunks did not fall within annotated genes. Low complexity regions were excluded from joint-calling, based on the DRAGEN hg38 blacklist regions. Chunks were later joined into chromosomes after variant filtering and annotation steps described below.

Variant Quality Score Recalibration (VQSR) was used according to the GATK Best Practices recommendations to filter jointly-called variants to keep true genetic variants and remove sequencing artifacts, resulting in a high-confidence cohort of variants (19, 20). Variants exhibiting excess heterozygosity were labeled using variant filtration, and variants with > 54.69 scores were excluded from further analysis. We used VariantRecalibrator to calculate VQSLOD tranches for INDELs and SNPs using the recommended population resources and priors including dbSNP138 (21), 1000genomes gold standard INDELs (22), 1000genomes phase 1 high confidence SNPs (23), and Hapmap 3.3 SNPs (24). Specifically, our pipeline followed the parameters and steps listed in the 1.1.1 version of the “broad-prod-wgs-germline-snps-indels” pipeline. This pipeline provided VariantRecalibrator with several per-site metrics from the NBB cohort, including depth of coverage, mapping quality, Fisher strand, rank sum test for relative positioning, and strand odds ratio. The resulting INDEL and SNP models were used to conduct score recalibration genome-wide.

Variants were annotated by ANNOVAR using data from refGene and dbSNP150 (21, 25, 26). The final cohort VCF contained 162,836,196 jointly-called variants. Genomic ancestry was broadly mapped to one of five 1000genomes reference panel super-populations using Peddy (27). Principal component analysis of population structure was performed on 220,819 autosomal variants from the VQSR filtered cohort VCF (MAF 5%, linkage disequilibrium pruned at 0.2) using SNPrelate (28).

### Huntington’s Disease association analysis

Variants were reduced to variants at or below 5% minor allele frequency (MAF) and those having a non-silent consequence on the protein sequence comprising: mis-sense, frameshift, stopgain, stoploss, or within the splice site of coding exons. Gene-collapsed optimal sequence kernel association testing (SKAT-O) and single variant testing was performed using EPACTS (https://github.com/statgen/EPACTS). Single variant testing and gene-collapsed testing was performed using covariates for biological sex, age, and the first 10 principal components. For single variant testing, tests were limited to non-silent variants with an allele frequency less than 0.05 and a minor allele count of 10 or greater. In addition, SKAT-O testing was restricted to genes with at least 3 different rare non-silent variants. The positive controls were comprised of 615 clinically diagnosed Huntington’s disease cases. Samples not labeled as Huntington’s by any diagnosis method were used as negative controls. A genome-wide significance threshold of less than 5×10^−8^ was used for all tests.

## RESULTS

### Sequencing of a large whole-genome cohort of central nervous system tissue donors

We generated whole-genome sequencing data from post-mortem brain tissue donors collected at six NeuroBioBank tissue repositories, collected prior to the end of 2021, to median 32x depth of coverage across the genome **(Table 1, Supplemental Table 1)**. The cohort consisted of 9,543 individuals, some of whom were related, that passed all quality metrics from a starting set of 9,680 samples submitted for sequencing from the six BTRs (**Supplemental Figure 1**). All of the 9,543 WGS samples correctly matched their counterpart present in the genotyping experiment as described in the methods. Whole genome sequencing showed robust quality metrics, which were consistent across samples received from different BTRs **(Supplemental Table 1)**. The population structure of the NeuroBioBank cohort had populations of ancestry similar to the underlying demographics observed in the United States population (29). This includes individuals who have ancestry which broadly matches the super-populations of European, African, East Asian, South Asian, and Admixed American, with the predominate cluster within this cohort most similar to European ancestry.

**Table 1.** Cohort statistics for samples within the NeuroBioBank whole-genome sequencing cohort. Sample statistics are displayed for samples that passed QA thresholds and are binned according to site. Demographic super-populations are abbreviated for those most similar to ancestry from Europe (EUR-Like), Africa (AFR-Like), South Asia (SAS-Like), East Asian (EAS-Like), and the Americas (AMR-Like). Individuals who displayed admixed ancestry were not categorized by most-similar ancestry.

| Biobank | Miami | Pittsburgh | Mt.Sinai | Sepulveda | Maryland | Harvard | Total |
| --- | --- | --- | --- | --- | --- | --- | --- |
| Passing Samples (Male) | 820 (469) | 572 (399) | 1575 (760) | 2419 (1245) | 1432 (824) | 2725 (1480) | 9543 (5177) |
| Mean Age (SD) | 65.6 (19.4) | 46.9 (13.7) | 76.2 (13.6) | 67.6 (17.3) | 41.2 (30.7) | 67.4 (17.4) | 60.8 |
| EUR-like | 651 | 470 | 1185 | 2216 | 1136 | 2627 | 8285 |
| AFR-like | 87 | 84 | 202 | 45 | 170 | 21 | 609 |
| SAS-like | 3 | 2 | 7 | 4 | 8 | 2 | 26 |
| EAS-like | 4 | 1 | 9 | 22 | 9 | 5 | 50 |
| AMR-like | 55 | 9 | 140 | 100 | 85 | 40 | 429 |
| Admixed | 20 | 6 | 32 | 32 | 24 | 30 | 144 |
| Total | 820 | 572 | 1575 | 2419 | 1432 | 2725 | 9543 |
| Avg Coverage Dragen | 37 (3.4) | 37.5 (4.0) | 37.3 (3.4) | 38.2 (4) | 37.4 (3.5) | 37.6 (4.2) | 37.5 |

Neurodegenerative disease (and, death, in general) occurs most frequently later in life and as a result older individuals are over-represented in the NeuroBioBank cohort (30). The average age is 60.8 across six BTRs. Four of the six BTRs skew towards older individuals, with an average age of 69.2, with samples collected at Mount Sinai being the oldest at 76.2 on average. Two BTRs, Maryland and Pittsburgh, historically have focused on collecting post-mortem brain tissue samples from younger individuals, with an average age of 41.2 and 46.9 respectively.

We used principal component analysis to assess genetic ancestry and population structure to help detect biases due to batching, sequencing, preparation and computational processing. After labeling individuals based on closest genetically-inferred super-population, pairs of principal components (PCs) were plotted up to PC6 **(Supplemental Figure 2 A-D)**. Plots revealed no PC clusters based on sequencing batch, repository, or other categories besides ancestry. A full listing of PCs 1-32 per sample are listed in Supplemental Table 1.

### Phenotypes of the NeuroBioBank cohort

There is a broad set of neurological and psychiatric phenotypes related to the brain tissues collected at each of the BTRs and paired with genotypes. These phenotypes are grouped into 15 different ICD disease categories. To better understand the distribution of these phenotypes, we tabulated the number of disease diagnoses and their ICD-10 codes (**Supplemental Table 2**). The disease diagnoses represented within the database are a one-to-many relationship, and one individual may have multiple unrelated diagnoses. For diagnoses not labeled by ICD-10 coding NBB custom labels were applied as described in the methods. These include NBB000 “no clinical brain diagnosis found” (*n=*1286) and NBB222 “diagnostic pathology not present” (*n=*2320). The overlap of these two labels is a set of 996 individuals who are coded with both “NBB000; no clinical brain diagnosis found” and “NBB222; diagnostic pathology not present”. Together this subset of individuals, collected and sequenced using the same methodology as individuals with diagnoses of brain disease, provides a high-quality sub-cohort absent of diagnosed brain-related disease.

**Table 2.** Counts by type of included and removed variants from the NeuroBioBank whole genome sequencing cohort joint calls.

|  |  |  |  |  |  |
| --- | --- | --- | --- | --- | --- |
| Total Variants | - | VQSR Strike | - | ExcessHet Strike | Passing Variants<br>(Total Variants - Strikes) |
| 206,429,754 |  | 34,798,225 |  | 510,320 | 171,121,209 |
| SNPs | + | Deletions | + | Insertions | Passing Variants<br>(SNP + Del + Ins) |
| 140,469,680 |  | 16,359,171 |  | 14,292,358 | 171,121,209 |
| Exonic | + | Intronic / UTR | + | Intergenic /<br>ncRNA | Passing Variants<br>(Exonic + Intronic + Intergenic) |
| 1,695,544 |  | 66,622,912 |  | 102,802,753 | 171,121,209 |
| Nonsynonymous<br>SNVs | + | Stop Loss / Gain<br>and Frameshift | + | Non-coding /<br>Synonymous | Passing Variants<br>(Non-silent + Silent) |
| 1,011,388 |  | 67,386 |  | 170,042,435 | 171,121,209 |

The most prevalent categories of disease within ICD-10 labeled clinical brain diagnoses are represented by codes G00-G99 “Diseases of the nervous system”, and F00-F99 “Mental, Behavioral and Neurodevelopmental disorders”. G codes accounts for 6312 individual clinical brain diagnoses, and F codes account for 4952 individual clinical brain diagnoses (**Figure 1A**). Other prevalent categories of disease include I codes “Diseases of the circulatory system” with 471 diagnoses, and Q codes “Congenital malformations, deformations and chromosomal abnormalities” with 226 clinical diagnoses. The exact number of diagnoses for a disease or category of disease presented in Figure 1 are not definitive, as updates to the NeuroBioBank phenotype portal are ongoing and new donors and new diagnoses to existing donors may be added.

**Figure 1.**
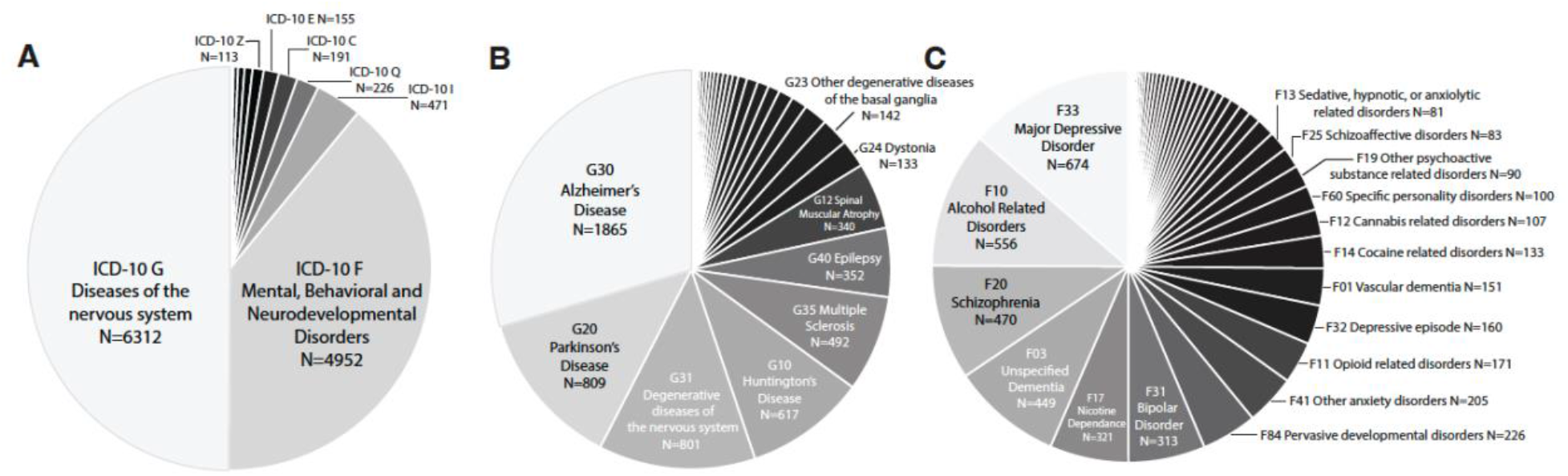
Disease distribution in the NeuroBioBank cohort for clinical diagnoses. A. Plot showing the most prevalent ICD-10 codes among clinical diagnoses, highlighting codes G and F as the primary components of the NeuroBioBank cohort. B. Prevalence of codes listing individual diseases and disease categories from ICD-10 G, diseases of the nervous system. C. Prevalence of codes listing individual diseases and disease categories from ICD-10 F, mental, behavioral, and neurodevelopmental disorders. Total number of diagnoses for each ICD-10 code are listed for each bin for the major components of each category.

The top three most prevalent clinical brain diagnoses among individuals within the NBB collection fall within the ICD-10 “G” category, diseases of the nervous system, and are of the neurodegenerative type. This includes Alzheimer’s disease (G30 *n=*1865), Parkinson’s disease (G20 *n=*809), and code G31 *n=*801 which encompasses “Other degenerative diseases” but is primarily composed of G31.0 Frontotemporal dementia (**Figure 1B**). Clinical diagnosis of Huntington’s disease (G10) was also highly prevalent with *n=*617 cases and is often accompanied by a corresponding genetic diagnosis. Other notable neurological diseases with more than 50 cases within the cohort are multiple sclerosis (G35; *n=*492), Epilepsy (G40; *n=*352), dystonia (G24; N=143), and migraine (G43; *n=*81).

The ICD-10 F category encompassing mental, behavioral, and neurodevelopmental disorders, is the second most prevalent set of diagnoses. Major depressive disorder (MDD; F33) was found to be the most commonly observed disorder among the F category of diseases (*n=*674) (**Figure 1C**). Other psychiatric diseases, such as schizophrenia (F20; *n=*470), and bipolar disorder (F31; *n=*313) were also highly represented in this cohort. Developmental disorders (F84) such as autism spectrum disorder (F84.0; *n=*120) and Rett’s syndrome (F84.2; *n=*101) are less common compared to MDD and schizophrenia, but still number in the hundreds of cases within the cohort. Lastly, in the ICD-10 F category there is a diverse set of substance abuse disorder diagnoses represented, with sample sizes ranging from a few dozen to hundreds, including opioid, nicotine, alcohol, and other substances of abuse.

Other notable prevalent diseases outside of the G and F ICD categories include cerebral infarction (I63 *n*=194), malignant neoplasm of the brain (C71; *n=*172), and sphingolipid disorders (E75; *n=*116). These three examples highlight the diversity of other nervous system disorders represented in the cohort, including metabolic disorders that impact the nervous system, congenital malformations which alter normal brain anatomy, and diseases whose origin is outside the nervous system but that can directly impact the brain. Within the phenotype database are multiple other diseases which can result in mortality that results in eligibility for brain donation and may only indirectly impact the brain. While the NBB sources data from central nervous system tissue, the recorded life histories and whole-genome nature of this data may record systemic conditions which is available for use by researchers.

### Variation in the NeuroBioBank cohort

To measure associations between phenotype and genetics, variants must be jointly called, filtered, annotated and combined into a cohort VCF. Specifically, this cohort VCF is an aggregation of the individual DRAGEN-produced gVCFs into one combined file, available per-chromosome from the NIMH Data Archive. After combining and jointly calling variants across the subset of 9,543 included samples, there were a total of 206,429,754 called variants (**Table 2**). Variants with excess heterozygosity were removed prior to variant recalibration. After filtering variants by Variant Quality Score Recalibration, the total number of variants was reduced to a passing set of 171,121,209 SNPs and INDELs across all autosomes, and chromosomes X, Y and M. A total of 35,308,545 SNPs and INDELs were identified as low quality and excluded from further analysis. The final set of 171,121,209 passing variants included 140,469,680 SNPs, 16,359,171 short deletions and 14,292,358 short insertions. This passing variant count is comparable to other studies with similar sample size and demographic characteristics (31). The concordance rate between WGS and genotyping array data was calculated to be 99.7% across 325,246,554 genotypes assessed.

After annotation, we identified the number of exonic variants as 1,695,544 SNPs, insertions and deletions, with exonic being defined as any genomic element falling within the borders of an annotated exon and excluding UTRs, introns, and intergenic regions. This set of exonic SNPs and INDELs included thousands of non-silent sites that have the potential to disrupt protein function and alter phenotype. This included 1,011,388 nonsynonymous mutations, 27,259 frameshift deletions, 13,271 frameshift insertions, 25,773 stop-gain mutations, and 1083 stop-loss mutations. While these non-silent changes are the most likely cause of changes in phenotype, the cohort also contains 170,042,435 non-exonic variants which may also contribute to disease. Together there is a wealth of high-quality variants, both within and between genes.

### Association of genes and variants to Huntington’s disease

To evaluate how successful the multi-institute pipeline of tissue collection, sequencing, and phenotype data collection was in producing associations between genetics and disease, we conducted a positive control association experiment to confirm the ability to detect robust links between sequence and neuropsychiatric disease. We tested records of individuals clinically diagnosed or clinically suspected of having Huntington’s disease (*N*=615), to replicate the known association of disease to repeat expansions in the *HTT* gene and its genetic markers. Of 620 individuals with Huntington’s disease in the NBB database, 617 had a clinical diagnosis of Huntington’s disease, and 615 had WGS available for testing. We compared these cases to a set of 8920 individuals with no history or no suspected history of Huntington’s disease. Exact case/control counts may differ depending on inclusion criteria per site tested. Using both the optimized Sequence Kernel Association Test (SKAT-O) and the single variant test as implemented in EPACTS using b.score, we tested genome-wide for association to the Huntington’s Disease phenotype.

The single variant association test identified a total of 12 loci that exceeded the Bonferroni corrected genome-wide significance threshold of p < 5×10^−8^, with the top 4 sites within the *HTT* gene on chromosome 4. The top 4 associated markers are large CAG repeats called as insertions into the *HTT* gene at the previously identified pathogenic site. These insertions range from 22 – 24 additional repeats added to the 21 CAG repeats normally present in the hg38 reference genome. Huntington’s disease patients typically have 40-50 repeats (32). The top site which contained an insertion of a 23 CAG repeats listed had a nominal p-value of 2.26×10^−109^ with the next highest and overlapping site having a nominal p-value of 4.61×10^−102^. Together the top 4 sites, plus a SNP in the nearby *UVSSA* gene, form a peak around the *HTT* gene (**Figure 2**). The remaining 7 loci outside of chromosome 4 which reached genome-wide significance were only marginally significant compared to the strength of association seen on chromosome 4. The *HTT* gene was found to be significantly associated with Huntington’s disease using the SKAT-O test with a nominal p-value of 8.51×10^−147^. The next nearest gene (*RGS12*), and only other SKAT-O significant gene, was directly adjacent to *HTT*, and reached significance due to the strength of association seen in *HTT* (**Supplemental Table 3**). Limiting the controls to *N*= 986 samples categorized as both NBB000 and NBB222 which describe individuals with no clinical brain or neuropathologic diagnoses, showed similar results with four single variants within the *HTT* repeat region being significant p < 5×10^−8^ and the *HTT* gene itself identified by SKAT-O with p = 1.66×10^−44^. Together, these results show an extremely robust positive association to Huntington’s disease, using both assumed Huntington’s-negative controls and controls with no neurological or psychiatric diagnosis, validating the quality and accuracy of the cohort sample collection, sequencing, and analysis.

**Figure 2.**
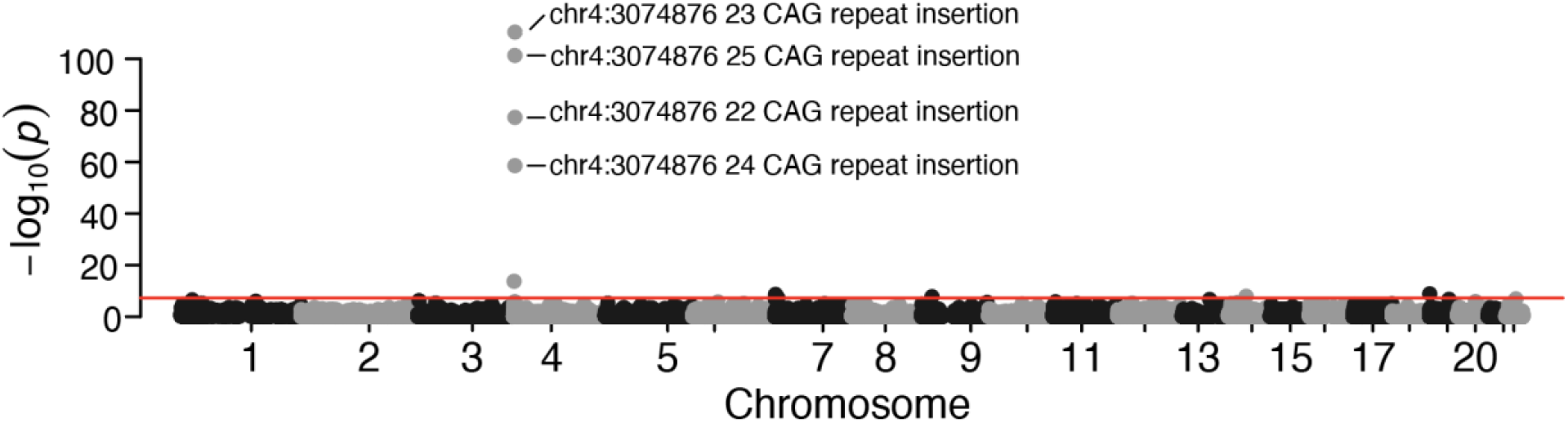
Manhattan plot of a rare single-variant genome-wide association to Huntington’s disease within the NeuroBioBank cohort VCF. Plotted are all exonic variants with an allele frequency between 0.0001 and 0.05. Huntington’s cases were defined as any individual with and ICD-10 clinical diagnosis of Huntington’s disease *N*=615. Individuals without a diagnosis of Huntington’s Disease were used as controls *N*=8920. The peak observed on chromosome 4 is centered on the *HTT* gene and the most associated site is the repeat expansion implicated in Huntington’s disease.

## DISCUSSION

The National Institutes of Health (NIH) NeuroBioBank repository is a combined effort of six brain tissue repositories to curate clinical data and biological samples to provide a unique resource to researchers who wish to advance knowledge about central nervous system diseases/disorders. Here we expand this resource by adding array genotypes and whole-genome sequencing data from a large cohort of high-quality samples and pair that with an extensive database of disease phenotypes. This data is made available to qualified researchers for a broad range of investigations into nervous system disease through the application process described on the NeuroBioBank website (https://nda.nih.gov/nda/access-data-info).

Collected from sites located across the United States, the NBB cohort contains a population of individuals that reflects the underlying demographics of that country. We acknowledge that the cohort is enriched for older individuals and individuals of European descent, but it also encompasses a wide variety of neurological diseases and psychiatric disorders. These diagnoses are focused on common brain-related disorders such as Alzheimer’s disease and related dementias, but the broad collection strategies of these BTR sites have also obtained many tissues diagnosed with rare clinical, neuropathologic and/or genetic diseases. We encourage researchers to explore the variety of diagnoses associated with tissue samples listed on the NeuroBioBank website. Included with the release of the NBB cohort are a variety of data types including the array-based genotype calls, and several single sample WGS result files: a per-sample genome-VCF, a structural variant VCF, CRAM alignment file, each with their associated metrics. In addition, a cohort VCF containing the filtered variants from 9,543 high-quality samples is available and accessible via the NIMH Data Archive.

The logistical challenges of any multi-site effort to sequence a large cohort may introduce sample swaps, duplicates, contaminations and other unknown sources of error. Evaluating for discrepancies in genetically determined biological sex and nearest ancestry showed results consistent with the self-reported values, when such self-reports were available. To demonstrate the quality and integrity of the WGS cohort for hypothesis testing, we conducted the validation experiment described above, that shows the reliability of our end-to-end workflow. Using a neurological disease with a well-described mechanism, Huntington’s disease, we were able to robustly identify known disease-causing repeats within the *HTT* gene using only genetic data and clinical diagnoses from this cohort. This validates the methodologies used at each step and supports future investigations of neurologic and psychiatric diseases. We hope that this reliable integration of genetic data with hundreds of disease diagnoses, and a network of bio-banked tissue, provides a robust resource for researchers for years to come.

While our study is comprehensive, it is crucial to acknowledge certain limitations inherent to this study. The nature of post-mortem brain tissue collection poses limitations as tissue represents a snapshot in time of the available population willing to donate brain tissue, resulting in biases in demography, disease etiology, and disease severity. In addition, environmental factors play a critical role in disease that whole-genome sequencing cannot capture. Our study and resource contribute to the broader understanding of the genetic aspects of central nervous system disorders, paired with tissue which can be used for complementary research methods to paint a more complete picture of each of those diseases.

It is also noteworthy that the WGS data described here represents a snapshot of the available donor samples collected at the time of sequencing. The NIH NBB is an ongoing effort, and the BTRs have banked additional tissue samples which are not included in this study but are available to researchers. It is also important to note that the study cohort described here does not represent an epidemiologically valid population study since each of the contributing brain banks has a history of special disease emphasis where different diseases are disproportionately represented in their collections.

The goal of this deep whole-genome sequencing project is to accelerate discovery of biological features and mechanisms which impact neurological and psychiatric diseases/disorders by providing a broad resource to the research community. By making available both the sequence, and the paired brain-tissue from which each sample was drawn, we hope to simplify access to the variation and material that will uncover the mechanisms that cause neuropathological and psychiatric diseases/disorders.

## Data Availability

Data from this study is available in the NIMH Data Archive, accessible at the following URL (https://nda.nih.gov/edit_collection.html?id=3917). To access, search and analyze this dataset, apply for access using the following URL (https://nda.nih.gov/nda/access-data-info). Tutorials further describing data access are available at the following URL (https://nda.nih.gov/nda/webinars-and-tutorials#tutorials). For further questions about data access, contact.

## Acknowledgements

We extend our heartfelt gratitude to individuals and their families who contributed brain tissue samples to this study. Processing of tissue specimens at the University of Miami for genotyping and WGS was supported by HHS contract 75N95019C00050. Tissue biobanking at University of Miami was supported by 75N95019C00050 and HHSN271201300028. Tissue biobanking at the University of Maryland was supported by contract numbers HHSN27520090011C, HHSN271201400045C, and 75N95019C00048. Tissue biobanking at the University of Pittsburgh was supported by contract number 75N95019C00047. Tissue biobanking at the University of California, Los Angeles was supported by contract number 75N95019C00045. Tissue biobanking at the Icahn School of Medicine at Mount Sinai was supported by contract number 75N95019C00049. Work at the Harvard Tissue Resource Center (HBTRC) was supported by contract number 75N95019C00046. We thank Jenny Edouard Pierre-Lys, Oluwarotimi Folorunso, Theresa Harvey, Kerenne Joseph, Anna Lally, Yuliya Leuchanka, Anil Vatsavayi, and Allison MacKenzie for their overall contributions to HBTRC. Sequencing and analysis at The American Genome Center, and the Center for Military Precision Health at Uniformed Services University was supported by contract numbers HU00012220044. Custom software used in these analyses is available upon request.

The views expressed in this manuscript are solely of the authors and do not reflect the official policy of the Departments of Army/Navy/Air Force, Department of Defense, USUHS, HJF, or the United States government. We report no conflicts of interest.

## Supplemental Material

**Supplemental Table 1 (Attached XLS)**. Detailed statistics about per sample performance for all samples within the NeuroBioBank cohort.

**Supplementary Table 2 (Attached XLS)**. Counts of ICD-10 categories observed in individuals included in the cohort and plotted in Figure 1.

**Supplementary Table 3 (Attached XLS)**. Results from testing the NeuroBioBank cohort for genetic associations to Huntington’s Disease based on 615 clinical diagnoses.

**Supplemental Figure 1.**
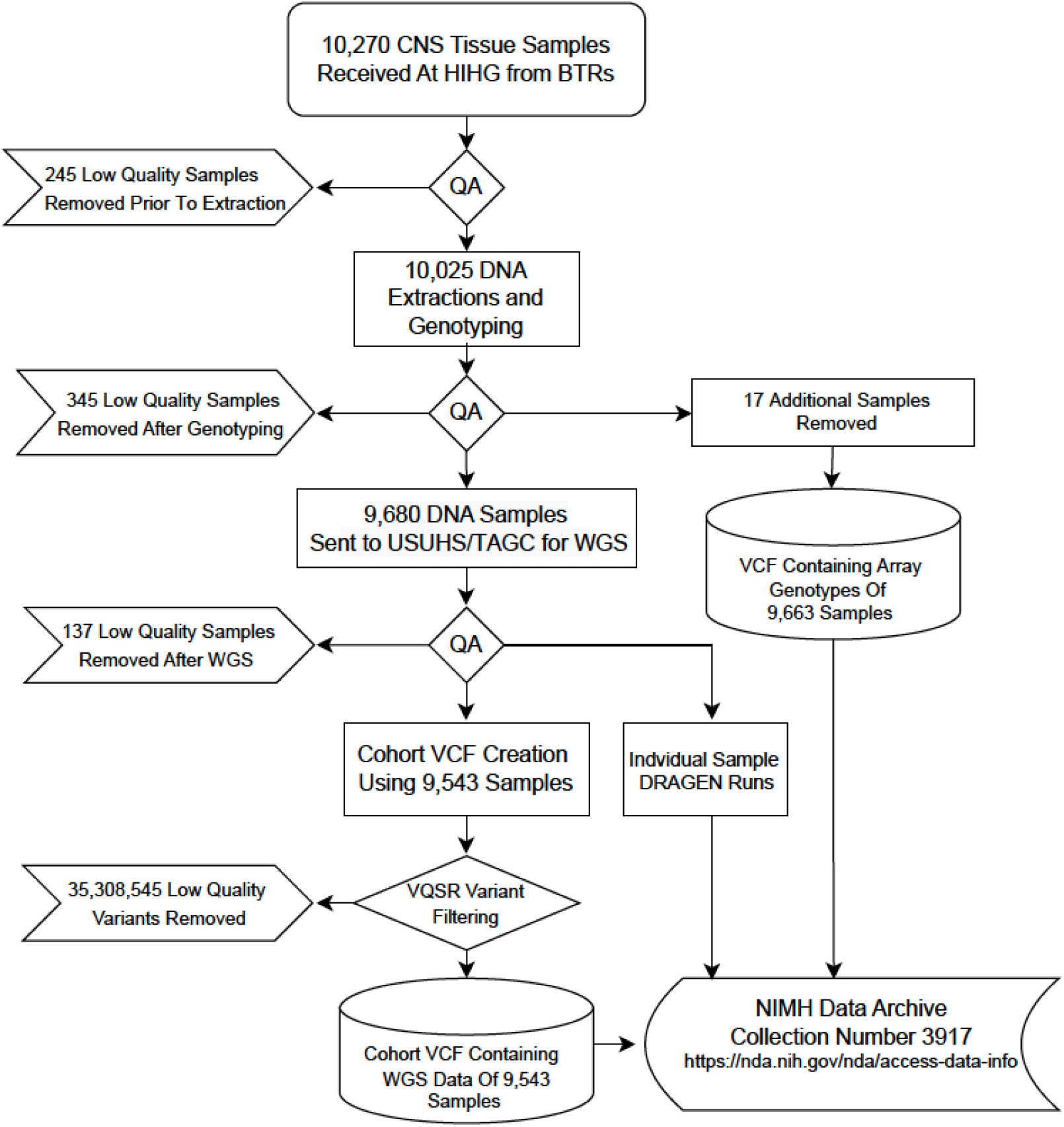
A flowchart of the sample collection, processing and analysis across six NeuroBioBank sites and two analysis working groups. Processes are placed within rectangle, decisions for adding/removing samples are within diamonds, removed sample counts are placed within flags, and databases and data storage are within cylinders. Final datasets can be found at the NDA URL under the collection number 3917.

**Supplementary Figure 2A.**
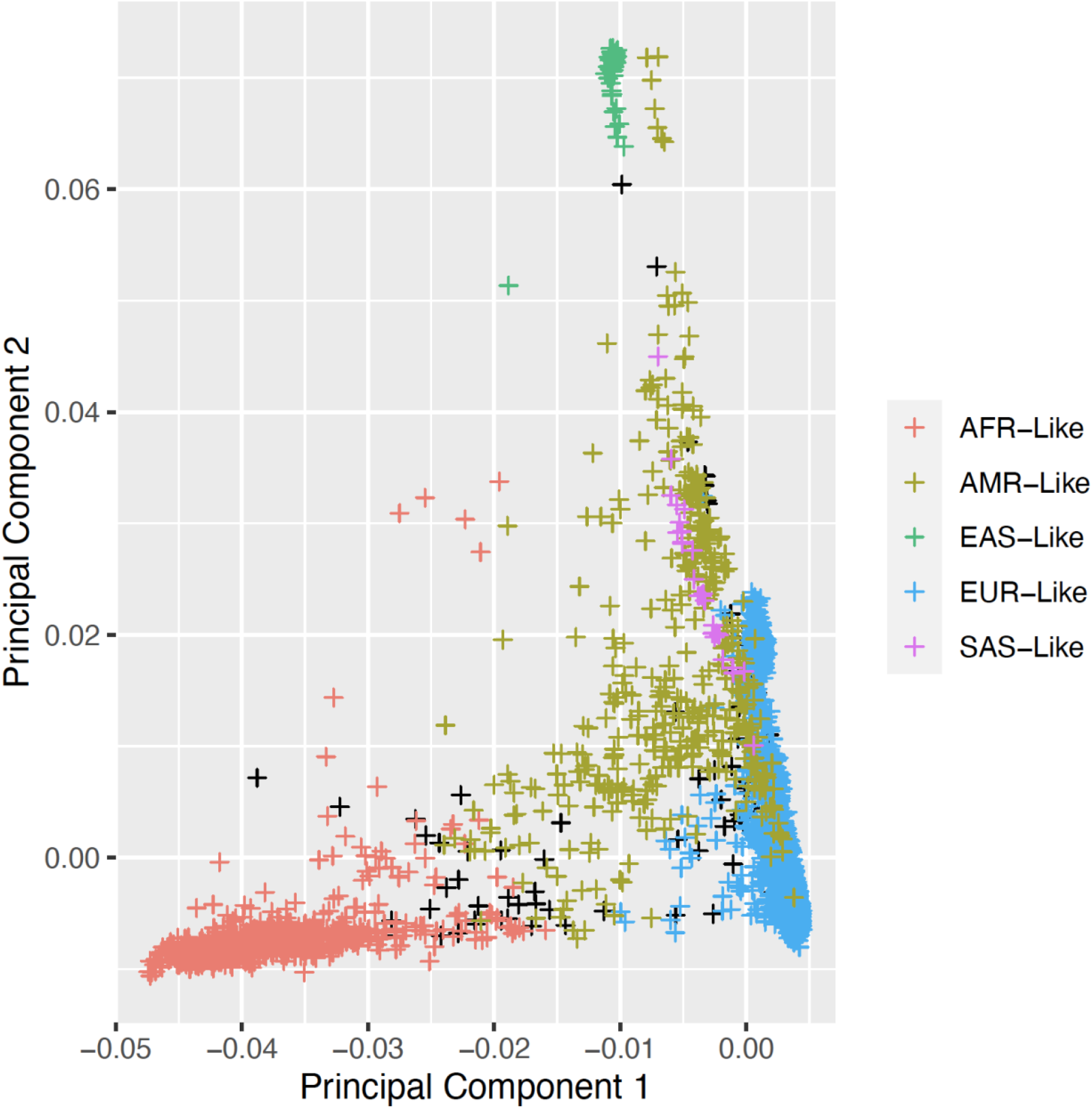
(Figures A-D in Attached PDF). Principal component analysis of the cohort of 9543 samples sequenced from the NeuroBioBank. Samples are colored by genetically inferred closest similarity to one of five 1000genomes superpopulations. These five categories consist of individuals most similar to a population of African descent (labeled as AFR-Like), European descent (EUR-Like), East Asian descent (EAS-Like), South Asian descent (SAS-Like), and admixed American (AMR-Like) as called by Peddy. Individuals where Peddy did not assign a superpopulation are colored in black. A. PCA of components 1 and 2. B. PCA of components 2 and 3. C. PCA of components 3 and 4. D. PCA of components 5 and 6

## References

1. Vaught J (2016): Biobanking Comes of Age: The Transition to Biospecimen Science. Annu Rev Pharmacol Toxicol. 56:211–228.

2. Freund M, Taylor A, Ng C, Little AR (2018): The NIH NeuroBioBank: creating opportunities for human brain research. Handbook of Clinical Neurology: Elsevier, pp 41–48.

3. Nichols L, Freund M, Ng C, Kau A, Parisi M, Taylor A, et al. (2014): The National Institutes of Health Neurobiobank: a federated national network of human brain and tissue repositories. Biol Psychiatry. 75:e21–22.

4. Prince M, Bryce R, Albanese E, Wimo A, Ribeiro W, Ferri CP (2013): The global prevalence of dementia: a systematic review and metaanalysis. Alzheimer’s & dementia. 9:63-75. e62.

5. Organization WH (2006): Neurological disorders: public health challenges. World Health Organization.

6. Heinzen EL, Neale BM, Traynelis SF, Allen AS, Goldstein DB (2015): The genetics of neuropsychiatric diseases: looking in and beyond the exome. Annual Review of Neuroscience. 38:47–68.

7. Kato T (2015): Whole genome/exome sequencing in mood and psychotic disorders. Psychiatry Clin Neurosci. 69:65–76.

8. Nicolas G, Charbonnier C, Campion D (2016): From Common to Rare Variants: The Genetic Component of Alzheimer Disease. Hum Hered. 81:129–141.

9. Funayama M, Nishioka K, Li Y, Hattori N (2023): Molecular genetics of Parkinson’s disease: Contributions and global trends. J Hum Genet. 68:125–130.

10. Cirulli ET, Goldstein DB (2010): Uncovering the roles of rare variants in common disease through whole-genome sequencing. Nat Rev Genet. 11:415–425.

11. Vornholt E, Luo D, Qiu W, McMichael GO, Liu Y, Gillespie N, et al. (2019): Postmortem brain tissue as an underutilized resource to study the molecular pathology of neuropsychiatric disorders across different ethnic populations. Neuroscience & Biobehavioral Reviews. 102:195–207.

12. Vialle RA, de Paiva Lopes K, Bennett DA, Crary JF, Raj T (2022): Integrating whole-genome sequencing with multi-omic data reveals the impact of structural variants on gene regulation in the human brain. Nat Neurosci. 25:504–514.

13. Hupalo D, Forsberg CW, Goldberg J, Kremen WS, Lyons MJ, Soltis AR, et al. (2022): Rare variant association study of veteran twin whole-genomes links severe depression with a nonsynonymous change in the neuronal gene BHLHE22. World J Biol Psychiatry. 23:295–306.

14. Wilkerson MD, Hupalo D, Gray JC, Zhang X, Wang J, Girgenti MJ, et al. (2023): Uncommon protein-coding variants associated with suicide attempt in a diverse sample of US Army soldiers. Biol Psychiatry.

15. Clark MM, Hildreth A, Batalov S, Ding Y, Chowdhury S, Watkins K, et al. (2019): Diagnosis of genetic diseases in seriously ill children by rapid whole-genome sequencing and automated phenotyping and interpretation. Sci Transl Med. 11.

16. Zhao S, Agafonov O, Azab A, Stokowy T, Hovig E (2020): Accuracy and efficiency of germline variant calling pipelines for human genome data. Sci Rep. 10:20222.

17. Goyal A, Kwon HJ, Lee K, Garg R, Yun SY, Kim YH, et al. (2017): Ultra-fast next generation human genome sequencing data processing using DRAGENTM bio-IT processor for precision medicine. Open Journal of Genetics. 7:9–19.

18. Jun G, Flickinger M, Hetrick KN, Romm JM, Doheny KF, Abecasis GR, et al. (2012): Detecting and estimating contamination of human DNA samples in sequencing and array-based genotype data. Am J Hum Genet. 91:839–848.

19. DePristo MA, Banks E, Poplin R, Garimella KV, Maguire JR, Hartl C, et al. (2011): A framework for variation discovery and genotyping using next-generation DNA sequencing data. Nature genetics. 43:491–498.

20. Van der Auwera GA, O’Connor BD (2020): Genomics in the cloud: using Docker, GATK, and WDL in Terra. O’Reilly Media.

21. Sherry ST, Ward M, Sirotkin K (1999): dbSNP—database for single nucleotide polymorphisms and other classes of minor genetic variation. Genome research. 9:677–679.

22. Mills RE, Luttig CT, Larkins CE, Beauchamp A, Tsui C, Pittard WS, et al. (2006): An initial map of insertion and deletion (INDEL) variation in the human genome. Genome research. 16:1182–1190.

23. Consortium GP (2012): An integrated map of genetic variation from 1,092 human genomes. Nature. 491:56.

24. Gibbs RA, Belmont JW, Hardenbol P, Willis TD, Yu F, Yang H, et al. (2003): The international HapMap project.

25. Wang K, Li M, Hakonarson H (2010): ANNOVAR: functional annotation of genetic variants from high-throughput sequencing data. Nucleic Acids Res. 38:e164.

26. O’Leary NA, Wright MW, Brister JR, Ciufo S, Haddad D, McVeigh R, et al. (2016): Reference sequence (RefSeq) database at NCBI: current status, taxonomic expansion, and functional annotation. Nucleic acids research. 44:D733–D745.

27. Pedersen BS, Quinlan AR (2017): Who’s Who? Detecting and Resolving Sample Anomalies in Human DNA Sequencing Studies with Peddy. Am J Hum Genet. 100:406–413.

28. Zheng X, Levine D, Shen J, Gogarten SM, Laurie C, Weir BS (2012): A high-performance computing toolset for relatedness and principal component analysis of SNP data. Bioinformatics. 28:3326–3328.

29. Colby SL, Ortman JM (2015): Projections of the Size and Composition of the US Population: 2014 to 2060. Population Estimates and Projections. Current Population Reports. P25-1143. US Census Bureau.

30. Hou Y, Dan X, Babbar M, Wei Y, Hasselbalch SG, Croteau DL, et al. (2019): Ageing as a risk factor for neurodegenerative disease. Nature Reviews Neurology. 15:565–581.

31. Telenti A, Pierce LC, Biggs WH, Di Iulio J, Wong EH, Fabani MM, et al. (2016): Deep sequencing of 10,000 human genomes. Proceedings of the National Academy of Sciences. 113:11901–11906.

32. Andrew SE, Paul Goldberg Y, Kremer B, Telenius H, Theilmann J, Adam S, et al. (1993): The relationship between trinucleotide (CAG) repeat length and clinical features of Huntington’s disease. Nature genetics. 4:398–403.

