## Supplementary figures and images for "The NeuroBioBank Whole-Genome Catalog: Sequencing from human brain donors with central nervous system disorders"

### Supplemental Figure 1

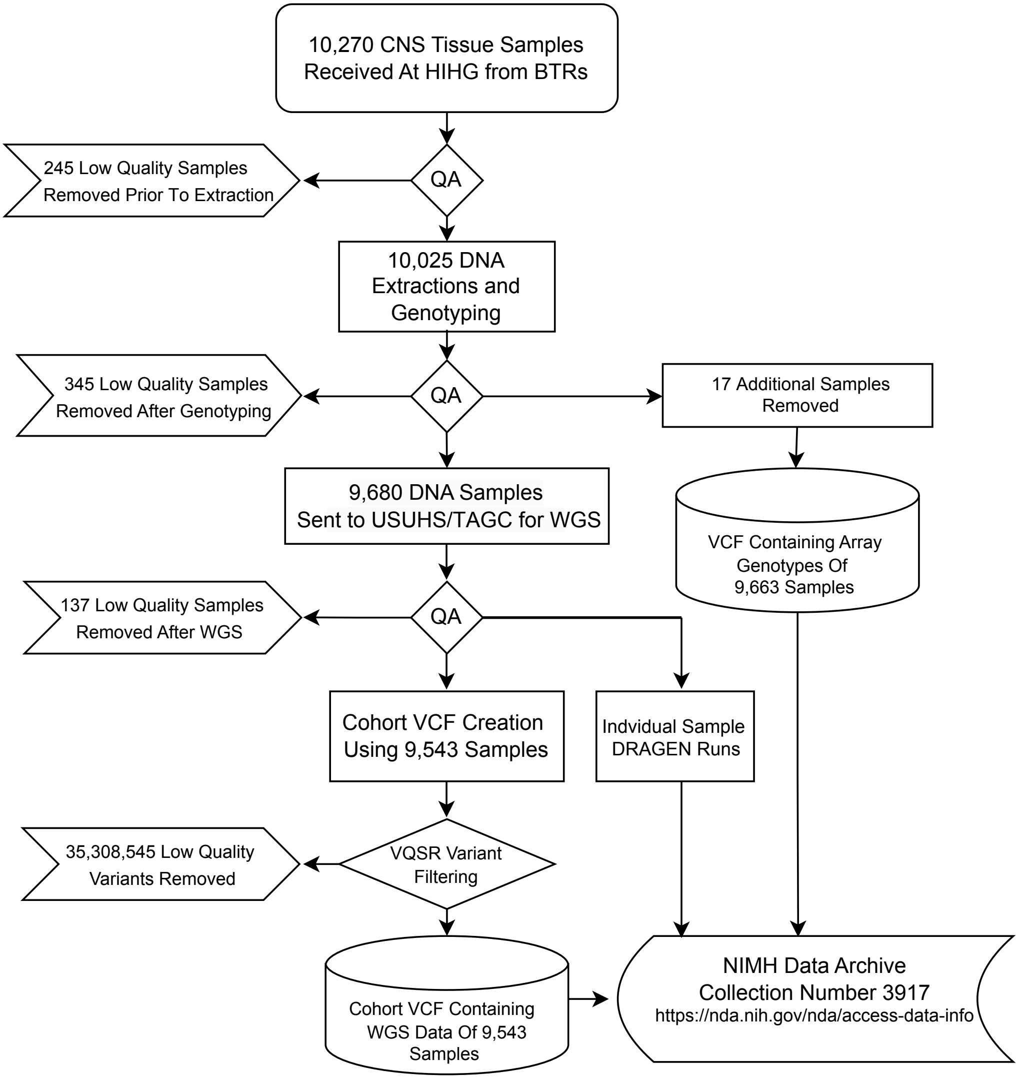

### Supplemental Figure 2 A-D

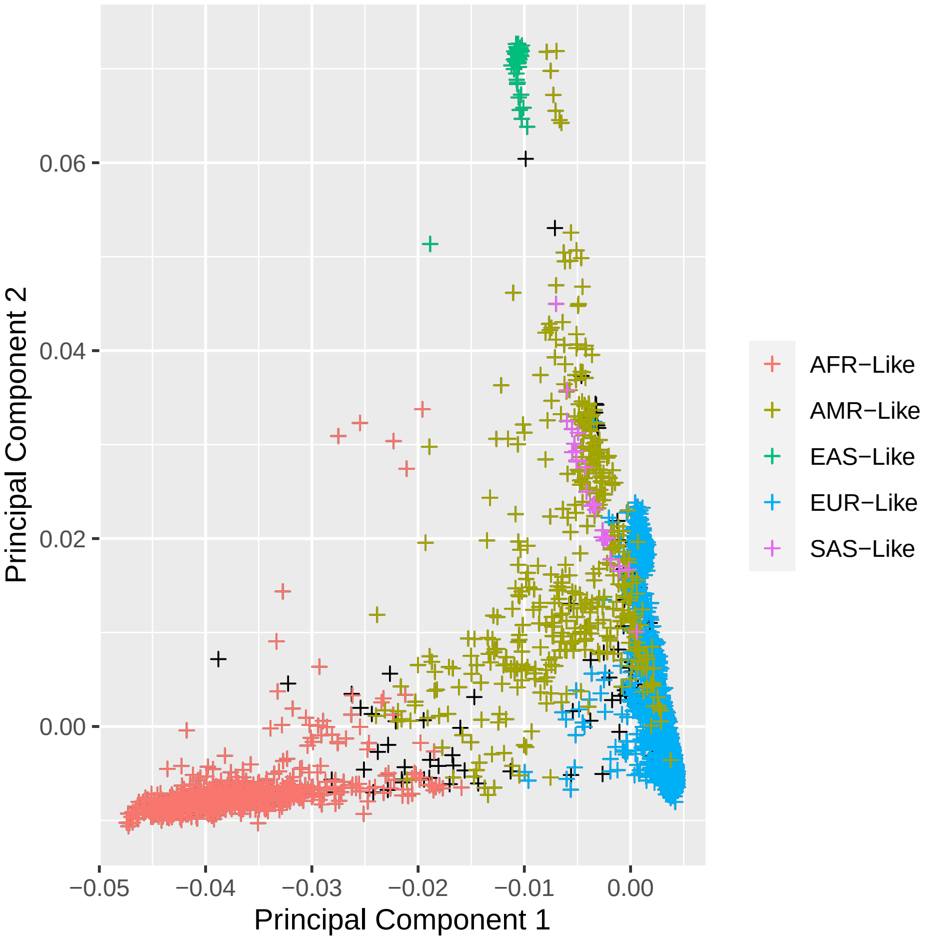

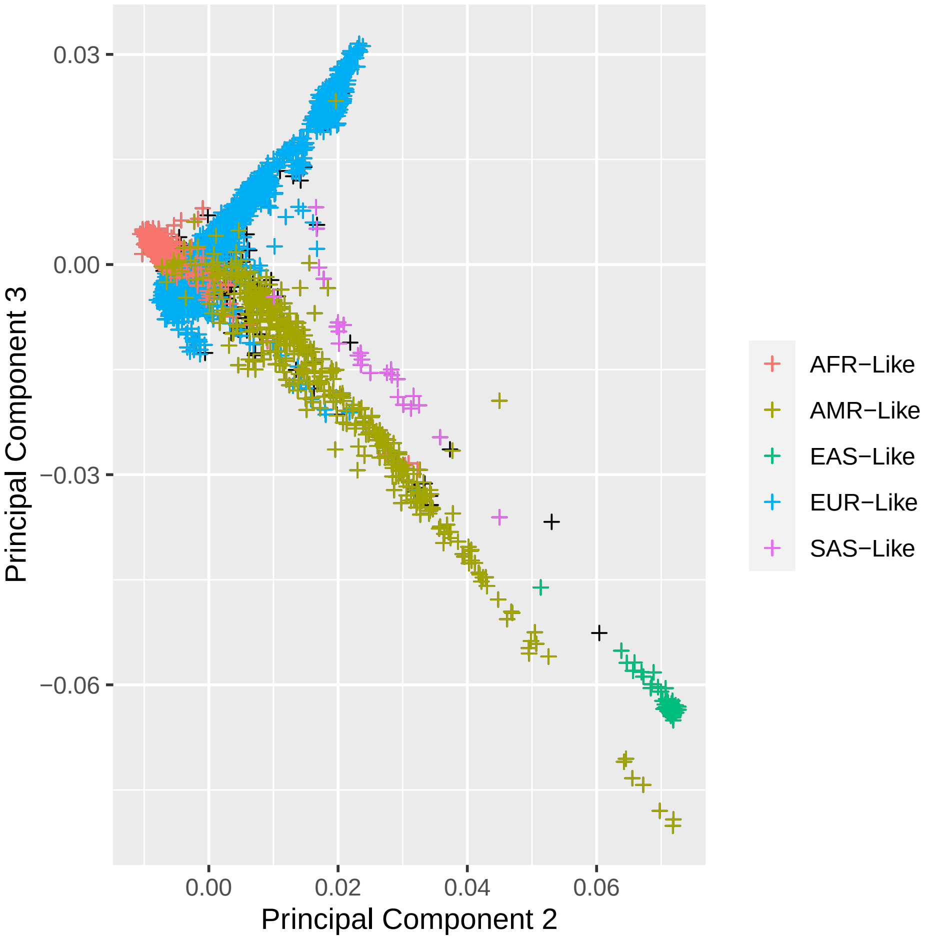

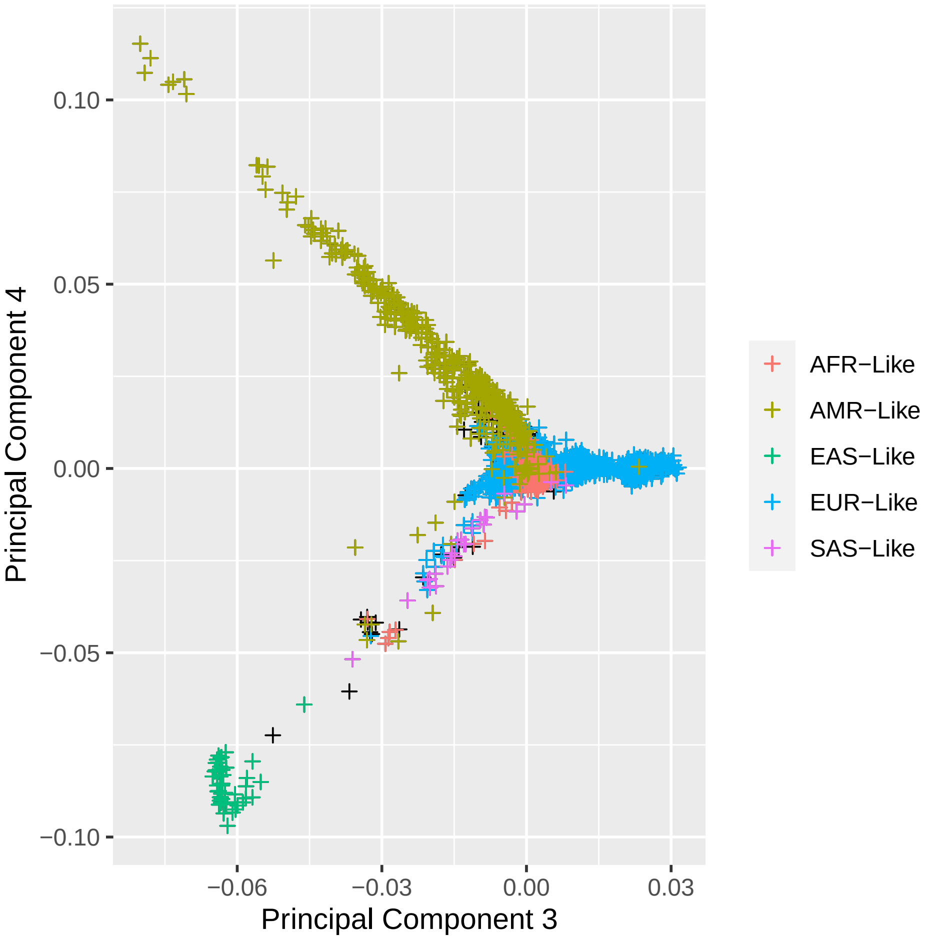

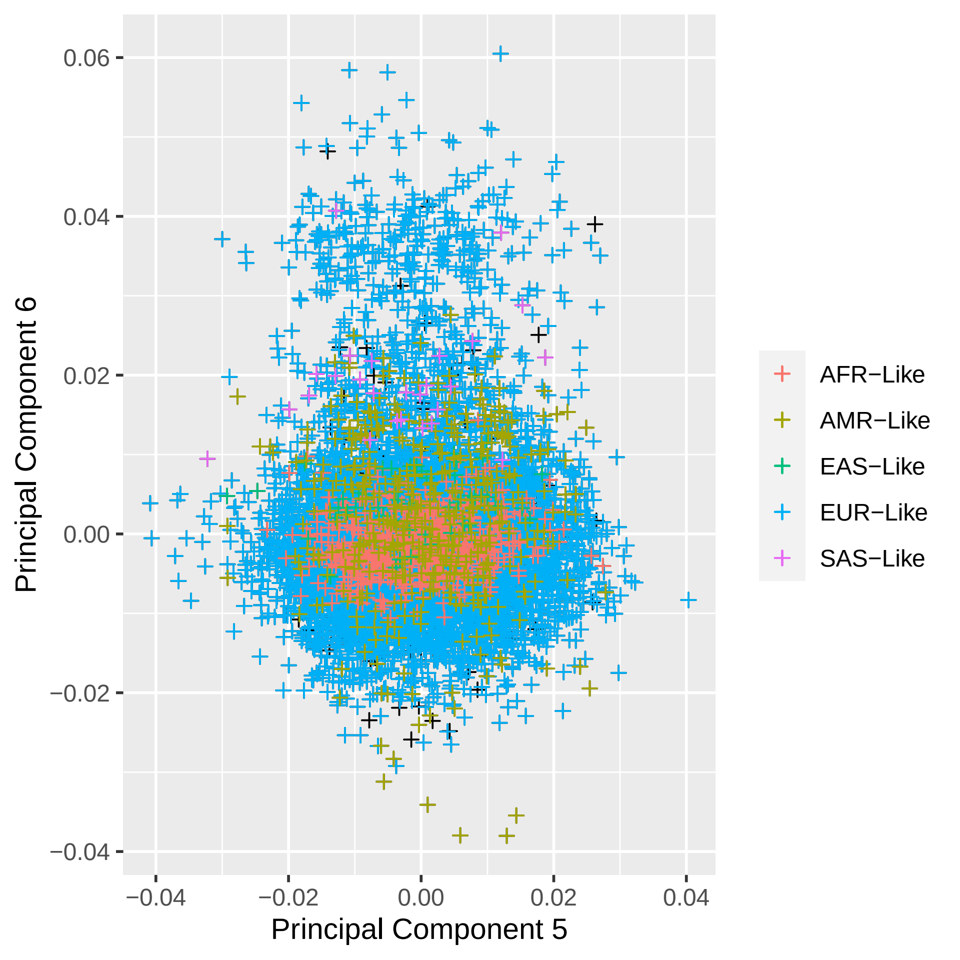
